# Repeated measures ASCA+ for analysis of longitudinal intervention studies with multivariate outcome data

**DOI:** 10.1101/2020.12.03.20243097

**Authors:** Torfinn S. Madssen, Guro F. Giskeødegård, Age K. Smilde, Johan A. Westerhuis

**Affiliations:** Department of Circulation and Medical Imaging, Norwegian University of Science and Technology, Trondheim, Norway; Biosystems Data Analysis Group, Swammerdam Institute for Life Sciences, University of Amsterdam, Amsterdam, The Netherlands

## Abstract

Longitudinal intervention studies with repeated measurements over time are an important type of experimental design in biomedical research. Due to the advent of “omics”-sciences (genomics, transcriptomics, proteomics, metabolomics), longitudinal studies generate increasingly multivariate outcome data. Analysis of such data must take both the longitudinal intervention structure and multivariate nature of the data into account. The ASCA+-framework combines general linear models with principal component analysis, and can be used to separate and visualize the multivariate effect of different experimental factors. However, this methodology has not yet been developed for the more complex designs often found in longitudinal intervention studies, which may be unbalanced, involve randomized interventions, and have substantial missing data. Here we describe a new methodology, repeated measures ASCA+ (RM-ASCA+), and show how it can be used to model metabolic changes over time, and compare metabolic changes between groups, in both randomized and non-randomized intervention studies. Tools for both visualization and model validation are discussed. This approach can facilitate easier interpretation of data from longitudinal clinical trials with multivariate outcomes.

**Author summary:** Clinical trials are increasingly generating large amounts of complex biological data. Examples can include measuring metabolism or gene expression in tissue or blood sampled repeatedly over the course of a treatment. In such cases, one might wish to compare changes in not one, but hundreds, or thousands of variables simultaneously. In order to effectively analyze such data, both the study design and the multivariate nature of the data should be considered during data analysis. ANOVA simultaneous component analysis+ (ASCA+) is a statistical method which combines general linear models with principal component analysis, and provides a way to separate and visualize the effects of different factors on complex biological data. In this work, we describe how repeated measures linear mixed models, a class of models commonly used when analyzing changes over time and treatment effects in longitudinal studies, can be used together with ASCA+ for analyzing clinical trials in a novel method called repeated measures-ASCA+ (RM-ASCA+).

## 1 Introduction

In recent decades, scientific and technological developments have increased our ability to both collect and manage large amounts of data. In biomedicine this has contributed to the rise of various “-omics”-fields (e.g. genomics, transcriptomics, proteomics, and metabolomics), where the focus is not on single variables such as individual genes, proteins or metabolites, but rather on the whole genome, proteome, or metabolome. Because “-omics”-datasets may contain hundreds to thousands of variables, considering each variable individually may be inefficient. In addition, “-omics”-data are multivariate by nature, and performing separate analyses for each variable does not take the interrelatedness between the variables into consideration.

A subset of the methods developed to analyze such data are based on the idea of combining two different kinds of statistical methods: 1) analysis of variance (ANOVA), and 2) dimensionality reduction methods such as principal component analysis (PCA) (1). Combining these methods in various ways allows the researcher to separate and visualize effects from different sources of variation in the data, while also accounting for the correlations between the outcome variables. One such method is ANOVA simultaneous component analysis (ASCA), where the response matrix is first decomposed into effect matrices according to the experimental design, and the impact of each experimental factor is visualized by applying PCA to the effect matrices (2). This methodology has since been extended to unbalanced designs by the adoption of the general linear model (GLM)-framework in estimating the effects, in a method called ASCA+. (3). A related approach was recently developed, termed linear mixed model-PCA (LiMM-PCA), in which the ASCA+ methodology is adapted to include random effects (4).

Longitudinal intervention studies with repeated measurements over time are an important type of experimental design in biomedical research. Such designs allow separating within-subject variation from between-subject variation, and also permit the study of trends over time. However, repeated measures data have properties which preclude the use of standard linear regression methods, such as fixed effects ANOVA, the most important one being that the observations belonging to the same individual are not independent. While the classical ASCA methodology has long been applied for longitudinal data analysis (2, 5), this approach has generally required strongly balanced designs to yield valid results. In contrast, longitudinal intervention studies routinely have many complicating features, such as unbalanced designs, randomized interventions, and substantial missing data. Repeated measures linear mixed models provide a powerful way to handle these issues (6), but these capabilities have so far not been extended to the multivariate case.

In this paper we introduce a new methodology, repeated measures ASCA+ (RM-ASCA+), using repeated measures linear mixed models in conjunction with ASCA+. We show how this method can be used in the analysis of longitudinal multivariate data with unbalanced designs and missing outcome data, including both visualization of results and assessment of model uncertainty. We also discuss other linear models for longitudinal data which can be used to estimate the effects, and comment on their suitability for ASCA+. We discuss how to adjust the analyses depending on whether the intervention is randomized or not, and we illustrate these differences by analyzing two different metabolomics datasets. We compare our findings with the original research papers for these datasets, and discuss the added benefit of RM-ASCA+ for biological interpretation in this setting.

## 2 Methods

### 2.1 Linear models for longitudinal data

A key step in ASCA+ is to define an appropriate linear model to use for estimating the effects. In this section we discuss some possible model types for analysis of longitudinal repeated measures data, and comment on their suitability for ASCA+. These models differ both in how they handle missing data, and whether they control for the baseline value of the response variable. The latter point can strongly affect, and even reverse, the effect estimates if the groups come from different pre-baseline populations, which is known as Lord’s paradox (7). The models to be discussed are: 1) repeated measures models, 2) longitudinal analysis of covariance (ANCOVA), and 3) analysis of changes. All model types discussed here involve including a random intercept for each subject, except for longitudinal ANCOVA and analysis of changes in the case of only two measurement occasions, as in this setting each subject only appears once in the response vector, and there is no need to account for within-subject correlation. The linear models presented here are the same as presented in the paper by Twisk et al., concerning different ways of analyzing randomized controlled trials with repeated measurements (8).

#### 2.1.1 Repeated measures

In a repeated measures model, the baseline value of the response variable is included in the responses in the same way as the follow-up measurements. Suppose that a response variable *y* is measured at *K* timepoints (*k* = 1..*K*) for *I* subjects (*i* = 1..*I*) belonging to one of *H* groups (*h* = 1..*H*). If the number of timepoints *K* = 3, and the number of groups *H* = 2, then a repeated measures model for this data is:

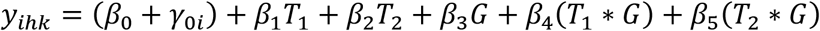

where *β*_*0-5*_ are the coefficients to be estimated from the data, *T*_*1*_ and *T*_*2*_ are indicator variables for the time factor, *G* is an indicator variable for the group factor, *T*_*1*_\**G* and *T*_*2*_\**G* are interaction variables, and *γ*_*0i*_ represents a subject-specific random intercept. Because the marginal mean for each timepoint is estimated individually, all subjects with at least one available response are included in the analysis. The indicator variables for time (*T*_*1*_ and *T*_*2*_) are either 0 or 1, i.e. they are reference coded with the first timepoint (baseline measurement) as the reference level. Depending on whether the indicator variable *G* is reference coded or sum coded, the coefficients for time, *β*_*1*_ and *β*_*2*_, will represent either the time effect for the reference group, or the time effect across both groups. In both cases *β*_*3*_ represents the group differences at baseline, while the interaction effects *β*_*4*_ and *β*_*5*_ represent the group differences in within-group change from baseline at each of the timepoints.

When assessing whether change in the response variable over time is different between the groups, a decision has to be made whether an adjustment for baseline differences in the response variable should be made. While adjustment also can be made for other baseline covariates, baseline adjustment will here refer to an adjustment for baseline differences in the response variable, unless otherwise stated. Although it is often believed that such an adjustment is made by assessing changes instead of directly comparing means, this is not correct (8). Suppose that change in the response variable is being compared between two groups, and that the true effect of both time and group is zero. Because of regression to the mean, the group with the highest baseline value will be expected to decrease slightly more than the group with the lowest baseline value (and vice versa), even if the treatment has no effect. Simply comparing changes can therefore lead to either over- or underestimation of the true treatment effect. The interaction coefficients *β*_*4*_ and *β*_*5*_ in the above model are therefore unadjusted, because they are only assessing whether the within-group change is different between the groups. Typically, one adjusts for a variable by including it as a covariate in a model. However, in this model, where the baseline values already are included in the responses, an adjustment can instead be made by removing the main effect for treatment, β_3_G, from the model while keeping its interactions with time:

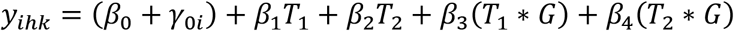

In this model, the estimated group means are constrained to be the same at baseline. This is also known as a constrained longitudinal data analysis (cLDA) model (9), while the previous model with the main effect for treatment included is sometimes referred to as an unconstrained longitudinal data analysis (ucLDA) model. The result of this constraint is that the interaction effects *β*_*3*_ and *β*_*4*_ will be adjusted for baseline.

The decision of whether to adjust for baseline depends primarily on the study design, and the research question of interest. In analysis of randomized controlled trials with a continuous response variable, adjusting for baseline is recommended, because it improves the precision of the treatment effect estimate (10, 11). In non-randomized studies, however, where uncontrolled group differences can exist prior to the study, adjustment for baseline can induce spurious effects if the groups originate from different pre-baseline populations (12). This phenomenon is known as Lord’s paradox, and implies that baseline adjustments must be done with care in non-randomized settings. In general, the decision to adjust for any covariate is dependent on prior knowledge about the study design and the measured variables, as well as on assumptions about how they causally interact. This applies to all forms of covariate adjustment, of which baseline adjustment is only a special case. Causal diagrams in the form of directed acyclic graphs provide a general framework for determining whether adjustment for a given variable creates or reduces bias with respect to the effect of interest (13).

#### 2.1.2 Longitudinal ANCOVA

The method of longitudinal ANCOVA involves using the baseline value, y_ih1,_ as a covariate, instead of modeling it as a response together with the follow-up values. This model can be written as:

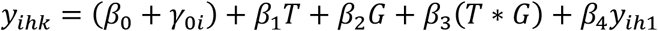

In this example, because the first timepoint is not included in the response vector, only two timepoints are represented in *y*, which can be described by one indicator variable *T*. In this model *β*_*2*_ represents the treatment effect at the first follow-up timepoint, while (*β*_*2*_ *+ β*_*3*_) represents the treatment effect at the second follow-up timepoint. Clearly, longitudinal ANCOVA also involves an adjustment for baseline, because it is included as a covariate in the model. It can be shown that cLDA and longitudinal ANCOVA are mathematically related; Differences in point estimates and confidence intervals for the group effect disappear with increasing sample size under randomization, and are usually small for non-randomized data (11). However, a possible disadvantage of longitudinal ANCOVA is that it cannot be used to model change from baseline. Another disadvantage is that subjects with missing baseline data are excluded from the analysis. The inclusion of the baseline response as a covariate also changes the meaning of the intercept term, possibly making the interpretation of the random intercept less intuitive.

#### 2.1.3 Analysis of changes

Analysis of changes involves expressing all follow-up values as differences from baseline, without including the baseline response as either a response or covariate in the model. An ANOVA, or linear model is then made to assess whether the average *Δ* differs significantly depending on group and time:

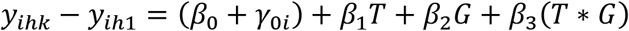

Testing *β*_*2*_ and (*β*_*2*_ *+ β*_*3*_) in this model effectively amounts to testing the same null hypotheses as *β*_*4*_ and *β*_*5*_, respectively, in the ucLDA-model, namely whether the time effect, or equivalently, the within-group change, is the same in both groups (11). One difference lies in how missing responses are handled. While all available response data is used in ucLDA, analysis of changes excludes responses where either the baseline or follow-up measurement is missing. As previously stated, this approach only assesses whether the change over time is significantly different depending on group, and a baseline adjustment is not made. If the baseline value is added as a covariate, then the model becomes equivalent to a longitudinal ANCOVA (8). Analysis of changes shares the previously mentioned disadvantages as longitudinal ANCOVA, such as not including the baseline measurements in the response vector, and poorer efficiency in the presence of missing outcome data.

### 2.2 ASCA^+^ / LiMM-PCA

#### 2.2.1 ASCA

In classical ASCA, the response matrix *Y* is decomposed into effect matrices according to the experimental design with standard ANOVA calculations, using differences in level averages.

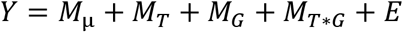

The resulting effect matrices are then analyzed and interpreted using PCA. However, this method is limited in that it only allows inclusion of fixed effects, and that classical ANOVA effect estimators based on differences in level means results in biased effect estimates for unbalanced designs. Classical ASCA is therefore only valid for longitudinal studies if the design is strongly balanced.

#### 2.2.2 ASCA+

In ASCA+, the original ASCA-methodology is extended to unbalanced designs by using general linear models (GLM) to estimate the effect matrices, instead of the classical ANOVA estimators based on differences in means. The GLM can be written as:

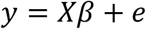

Where *y* is the response variable, *X* is the design matrix for the chosen linear model, *β* is the vector of coefficients to be estimated from the data, and *e* is a vector containing the residuals. In ASCA+, this equation is extended to multiple response variables.

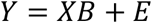

where *Y* now represents a response matrix with *J* columns (*j* = 1..*J*), and *B* represents a parameter matrix, where the *j*-th column of *B* corresponds to the regression coefficients belonging to the *j*-th column of *Y*. A GLM based on the design matrix *X* is then applied to every column of *Y*, and the coefficients are collected in the matrix *B*.

After estimation of the matrix *B*, the effect matrices are then made by multiplying the corresponding columns of *X* together with the corresponding rows of *B*. For example, in order to make the effect matrix for the time effect, all columns in *X* and rows in *B* except those belonging to the time factor are turned into zero, and the following operation is done to produce the effect matrix for time, *M*_*T*_:

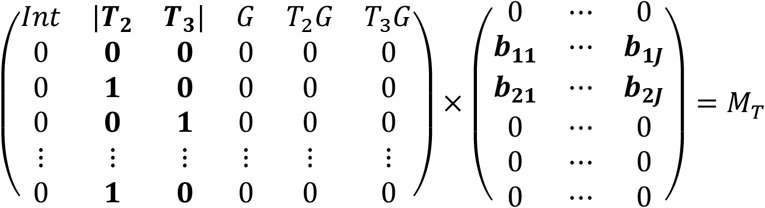

The effect matrix *M*_*T*_, which now contains the estimated level averages for the factor for time, can be analyzed by PCA in order to visualize the multivariate differences between the timepoints.

#### 2.2.3 LiMM-PCA

In LiMM-PCA, the ASCA+ methodology is further adapted to also include random effects, making it a potentially suitable method for correctly modeling the longitudinal structure of intervention studies. In LiMM-PCA, the response matrix *Y* is initially analyzed by PCA, and the resulting score matrix is used as input for the effect matrix estimation instead of *Y*. This is done because the methods for variance partitioning and statistical inference in LiMM-PCA rely on the response variables being orthogonal, which is ensured by analyzing the score matrix for *Y* instead of *Y* directly. By excluding components explaining minimal variation, one also limits the necessary number of variables to analyze, thus making it more feasible for analysis of high-dimensional data. During visualization, the effect matrices estimated from the *Y*-score matrix are analyzed with PCA, and then the resulting effect matrix loadings are multiplied by the original loadings for *Y* in order to back-transform the variables to their original variable space.

#### 2.2.4 RM-ASCA+

In the methodology presented here, RM-ASCA+, we combine repeated measures linear mixed models with ASCA+ to estimate the multivariate effects of time and the interaction between time and group in an unbalanced setting, while also accounting for within-subject dependency. We do this without applying the initial PCA-step as done in LiMM-PCA. The effects are estimated directly based on the full response matrix. This simplifies the methodology, and makes our proposed validation method, shown in the next section, more straightforward to implement. The linear mixed model for a single response with a random intercept for subject may be written as:

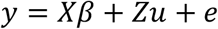

where *X* is the design matrix for the fixed effects according to the chosen linear model, *β* is a vector of fixed effect coefficients to be estimated from the data, *Z* is a dummy coded matrix for subject, and *u* is a vector containing the random intercepts. The fixed effects vector *β* is estimated by maximum likelihood methods, while *u* is predicted for each subject based on the estimated variance of *u*.

Like in ASCA+, this can be extended to multiple response variables.

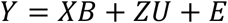

where *Y* now represents a response matrix with *J* columns (*j* = 1..*J*), and *B* represents a parameter matrix, where the *j*-th column of *B* corresponds to the regression coefficients belonging to the *j*-th column of *Y*, and *U* is a matrix containing the random intercepts. The effect matrices are then constructed as in ASCA+. If one wishes to visualize the joint impact of several experimental factors, it is possible to add effect matrices together by extending this operation out to more columns in *X* and rows in *B*:

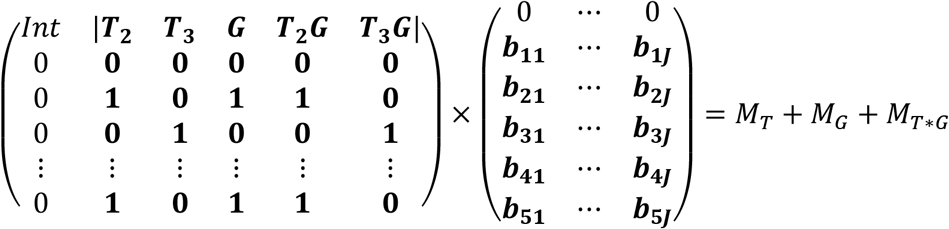

The resulting effect matrix, *M*_*T+G+T*\**G*_ contains the estimated level averages for all the combinations of the factor levels, and can be analyzed by PCA similar as *M*_*T*_. Although this approach confounds variation from different experimental factors, it may be useful during interpretation because it allows visualization the treatment effect in combination with the time effect.

In classical ASCA, the effect matrices are automatically mean-centered, because the mean matrix is subtracted from the data during the decomposition of *Y*, and the other effects are expressed relative to the grand mean. This does not happen with the coding system used here, where the factor for time is reference coded with baseline as reference. Instead, because the intercept represents the baseline mean, the rows in the matrix *M*_*T*_ belonging to baseline are set to zero. If *G* is sum-coded, then *M*_*G*_ *+ M*_*T*\**G*_ will be centered around zero, while if it is reference coded, the chosen reference group will have all its scores set to zero in *M*_*G*_ *+ M*_*T*\**G*_.

So far the random effects structure in the model is only used when estimating the fixed effects in *B*. However, the random effects themselves can also be included and visualized. The following method is adapted from a method by Zwanenburg et al., where variability between samples is visualized by projecting the effect matrix plus the residual matrix (e.g. *M*_*T+G+T*\**G*_ *+ E*), onto the effect matrix loadings (14). Similarly, we can also add the random effect matrix *ZU*, where *U* is the matrix containing the best linear unbiased predictions for the random effects for each variable, to the effect matrix together with the residuals, and project (*M*_*T+G+T*\**G*_ *+ ZU + E*) onto the loadings of the effect matrix *M*_*T+G+T*\**G*_. This allows visualizing the score trajectory of each individual in the loading space of *M*_*T+G+T*\**G*_, and is helpful for evaluating the size of the estimated effects relative to the between-subject variation and residuals.

#### 2.2.5 Model validation

In order to assess the robustness of the findings we use a seven-fold jackknife approach to assess variability in model parameters (15). This approach involves iteratively excluding a random subset of patients from the data, while keeping the proportion of groups constant, and then re-estimating the effects and the PCA-scores and loadings for the effect matrices. During each iteration the new loadings are rotated by orthogonal Procrustes rotation towards the original loadings, and the score matrix is rotated by multiplying it with the inverse of the resulting loading rotation matrix. The 2.5^th^ and 97.5^th^ percentiles of the jackknifed score- and loading estimates are then plotted as error bars around the estimates based on the full data. This approach only provides an approximate measure of the uncertainty of the estimated mean differences, and should not be interpreted as parametric 95 % confidence intervals. We mean center the effect matrices during every iteration, to avoid variability in the overall mean of the data to be incorporated in the error estimates.

### 2.3 Software and data analysis

All statistical analysis and figure creation were done in MATLAB 2019b. The fitlmematrix-function was used for mixed models, and the pca-function in the Statistics and Machine Learning Toolbox was used for PCA-analysis. MATLAB-code to reproduce the results is available on GitHub at (https://github.com/ntnu-mr-cancer/RM_ASCA).

## 3 Materials

To demonstrate RM-ASCA+, two published datasets are here used. These will be briefly described, and the reader is directed to their source publications for further details.

### 3.1 The NeoAva-trial

The first dataset used is from the NeoAva-trial, which is a randomized controlled trial assessing the effect of adding bevacizumab, an anti-angiogenic monoclonal antibody, to conventional chemotherapy in breast cancer patients with locally advanced HER2-negative tumors in a neoadjuvant treatment setting. In a study by Euceda et al., repeated tumor biopsies obtained over the course of treatment were analyzed with high resolution magic angle spinning (HR MAS) MR spectroscopy, using a CPMG sequence (16). The spectral region between 1.40 – 4.70 ppm, containing the majority of low-molecular weight metabolites, was selected as the region of interest, and spectral regions containing mostly lipids, ethanol, acetone and lidocaine were excluded. Spectra were PQN-normalized after removal of these areas (17), and metabolites were quantified by peak integration. For further details on spectral acquisition and processing we refer to the original publication by Euceda et al. (16). Metabolic changes were related to treatment group and tumor response. The dataset includes 16 quantified and log-transformed metabolites from 122 patients, of whom 60 received bevacizumab + chemotherapy, and 62 received chemotherapy only. Three tumor biopsies were taken; one before start of treatment, one after 12 weeks of treatment, and the last was taken from the surgically removed tumor. Data is missing at all timepoints, with 14 %, 36 %, and 29 % missing outcome data at each timepoint respectively, giving a total of 270 responses in the study.

### 3.2 Metabolic fingerprint after bariatric surgery

The second dataset used is from a study by Gralka et al., which prospectively assessed alterations in serum metabolites in patients undergoing one of three different kinds of bariatric surgery (proximal Roux-en-Y gastric bypass (RYGB), distal RYGB, and gastric sleeve) (18). Procedure selection was based on pre-existing clinical factors, such as degree of obesity, comorbidities, and patient preferences. Blood was drawn at baseline before surgery, and at 3, 6, 9, and 12 months after surgery, and the serum was analyzed using NMR spectroscopy. Spectra were obtained using a Bruker spectrometer operating at 14.1T, with a triple resonance inverse cryoprobe, and automatic tuning-matching unit and sample changer. A CPMG-sequence was used to acquire the spectra. The water region between 6.0 and 4.5 ppm was removed, and the non-normalized spectra were divided into 0.2 ppm bins, which were integrated using AMIX software (version 3.8.4; Bruker BioSpin). Thirty metabolites were quantified, of which 24 showed variability over time, and 21 were included in the published dataset. The study design is unbalanced, with 60 patients undergoing distal RYGB, 27 undergoing proximal RYGB, and 19 undergoing gastric sleeve. Data is missing at all post-baseline timepoints, with 7 %, 8 %, 12 %, and 32 % missing outcome data at each post-baseline timepoint, respectively, giving a total of 463 responses in the study. The metabolomics data was made freely available by the authors at the online repository MetaboLights, with the identifier MTBLS242.

## 4 Results

### 4.1 Effect of bevacizumab on tumor metabolism in breast cancer

As the NeoAva study is a randomized controlled trial, a constrained repeated measures model is used to estimate the effect matrices for RM-ASCA+. The following model was used:

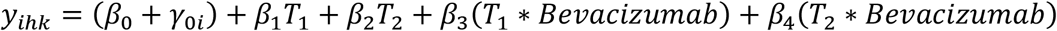

where the time factor was reference coded with baseline (*T*_*0*_) as reference, and the variable Bevacizumab was reference coded, where 1 indicates bevacizumab + conventional chemotherapy (Treatment), and 0 indicates conventional chemotherapy only (Control). In order to visualize how the treatment modified the metabolic changes during chemotherapy, the effect matrix for time and time*treatment interaction were added together, and the combined effect matrix was analyzed with PCA. While this confounds the variation from the time- and treatment factors, it also facilitates a more direct assessment of how the treatment and control groups differ at the different timepoints. The plots showing results for separate PCAs on each effect matrix are available in Supplementary Figure 1 and 2. All metabolite responses were log-transformed before analysis, and the effect matrix was mean-centered before PCA.

**Figure 1:**
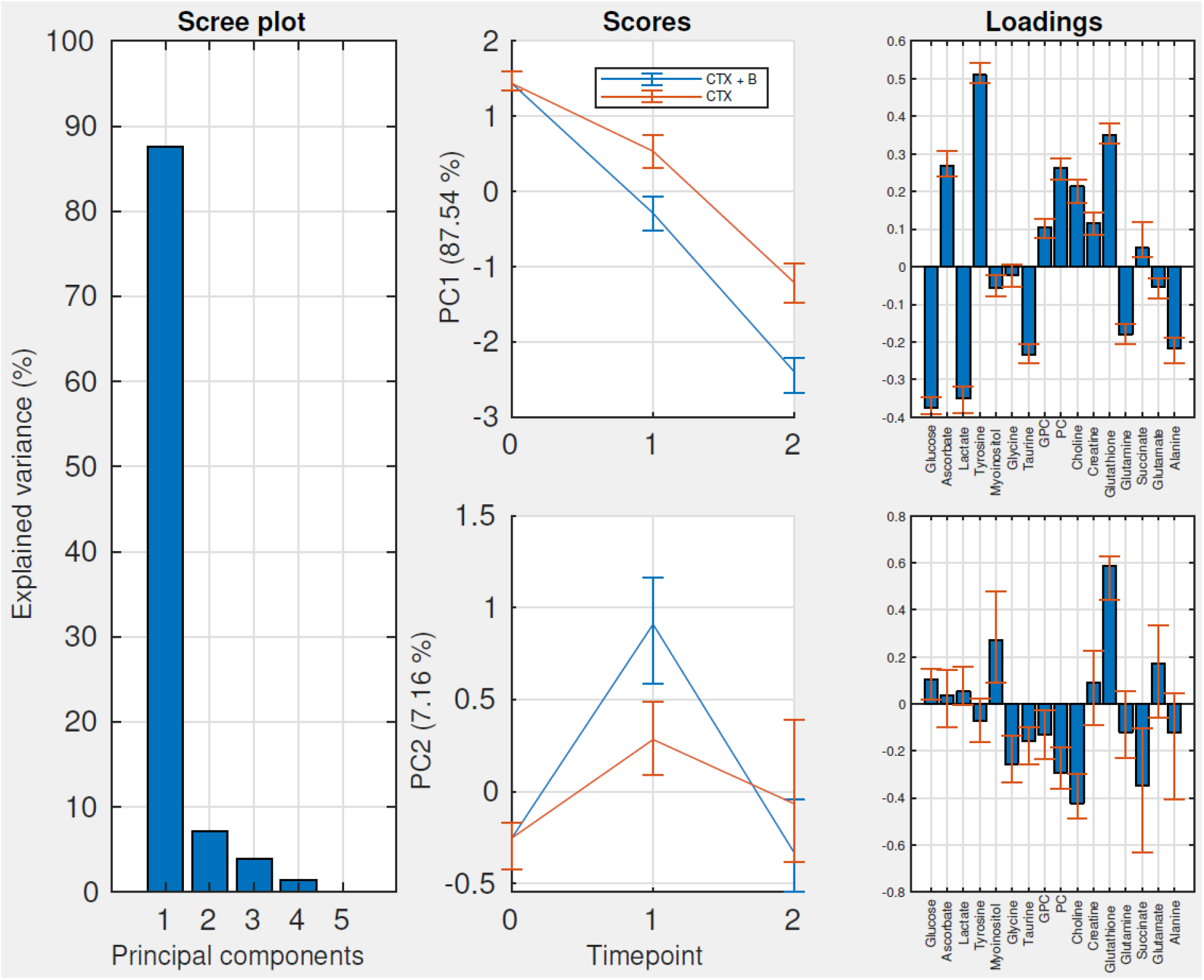
Scree-, score-, and loading plots for the full effect matrix for time + time*treatment. Abbreviations: PC: Principal component, CTX: Chemotherapy, B: Bevacizumab, GPC: Glycerophosphocholine, PC: Phosphocholine.

**Figure 2:**
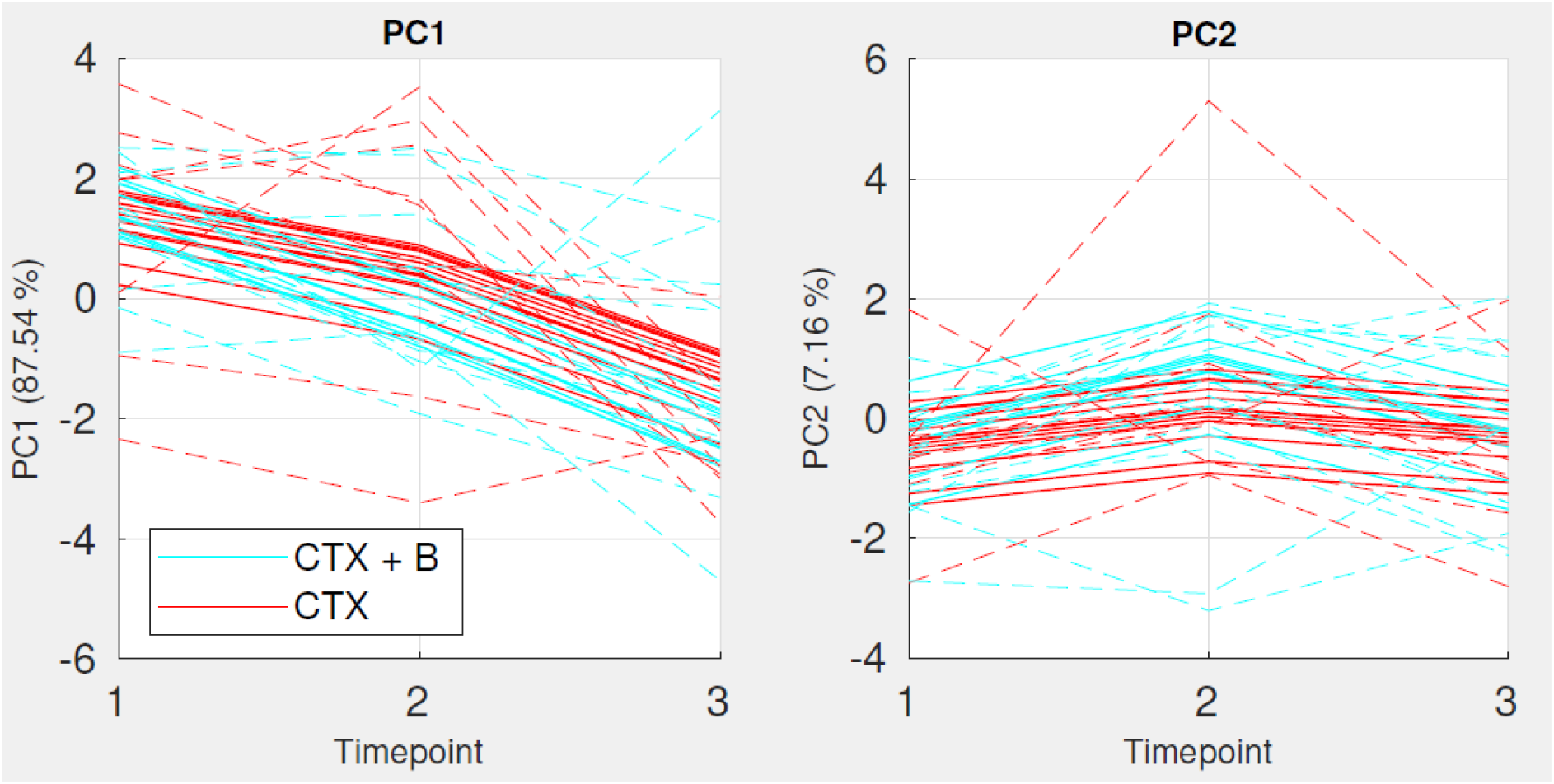
Score plots for PC1 and PC2 for the effect matrix for time + time*treatment, augmented with random effects (continuous lines) and both random effects and residuals (dashed lines). Only a random subset of patients with complete data are included in the plot.

The scree plot (Figure 1) shows that two principal components (PC) explain approximately 95% of the variation in the time and time*treatment interaction effect. The scores and loadings for PC1 show that levels of ascorbate, tyrosine, glycerophosphocholine, phosphocholine, choline, creatine and glutathione decrease over time, while levels of glucose, lactate, taurine, glutamine, and alanine increase over time, and that these changes are most rapid and pronounced for the treatment group.

In PC2, which explains only 7 % of the variation, both groups show an increased score value at the second timepoint compared with baseline, but the increase is higher in the treatment group. The PC2-loadings suggest that glutathione and myo-inositol are increased at this timepoint compared with the other timepoints, while glycine, phosphocholine, choline and succinate are decreased.

When the effect matrix for time + time*treatment is augmented with the random intercepts and residuals (Figure 2), we observe significant between-patient variation and heterogeneity in treatment response, showing that the estimated difference, while consistent, is modest relative to the variation between subjects and unexplained variation in the model. For comparison, we have projected both the fitted values (*M*_*T+G+T*\**G*_ *+ ZU*, continuous lines) and with the residuals added (*M*_*T+G+T*\**G*_ *+ ZU + E*, dashed lines), showing that there is significant residual variation after accounting for the between-subject variation.

### 4.2 Metabolic effects of bariatric surgery

To estimate the effects for RM-ASCA+ analysis of the bariatric surgery data, an unconstrained repeated measures model was used:

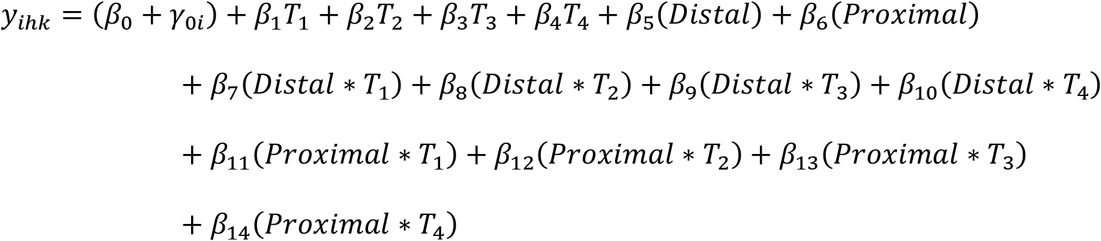

where time was reference coded with baseline as reference, and variables for treatment (Distal and Proximal) were sum coded, with the third category (Sleeve) specified by setting both Distal and Proximal equal to −1. With this coding system, the time effect represents the mean change over time across all groups, and the treatment and time*treatment interactions will be expressed as deviations from the time effect. In this analysis the effect matrices for the experimental factors are analyzed separately. To make the effect matrix for time, a matrix containing the coefficients *β*_*1*_*-β*_*4*_ for each metabolite was multiplied with their corresponding columns in the design matrix *X*. To make the effect matrix for (treatment + time*treatment), the same was done for the coefficients *β*_*5*_*-β*_*14*._ The results from the combined effect matrix is shown in Supplementary Figure 3. Before analysis every metabolite was square root transformed and scaled to its baseline standard deviation.

**Figure 3:**
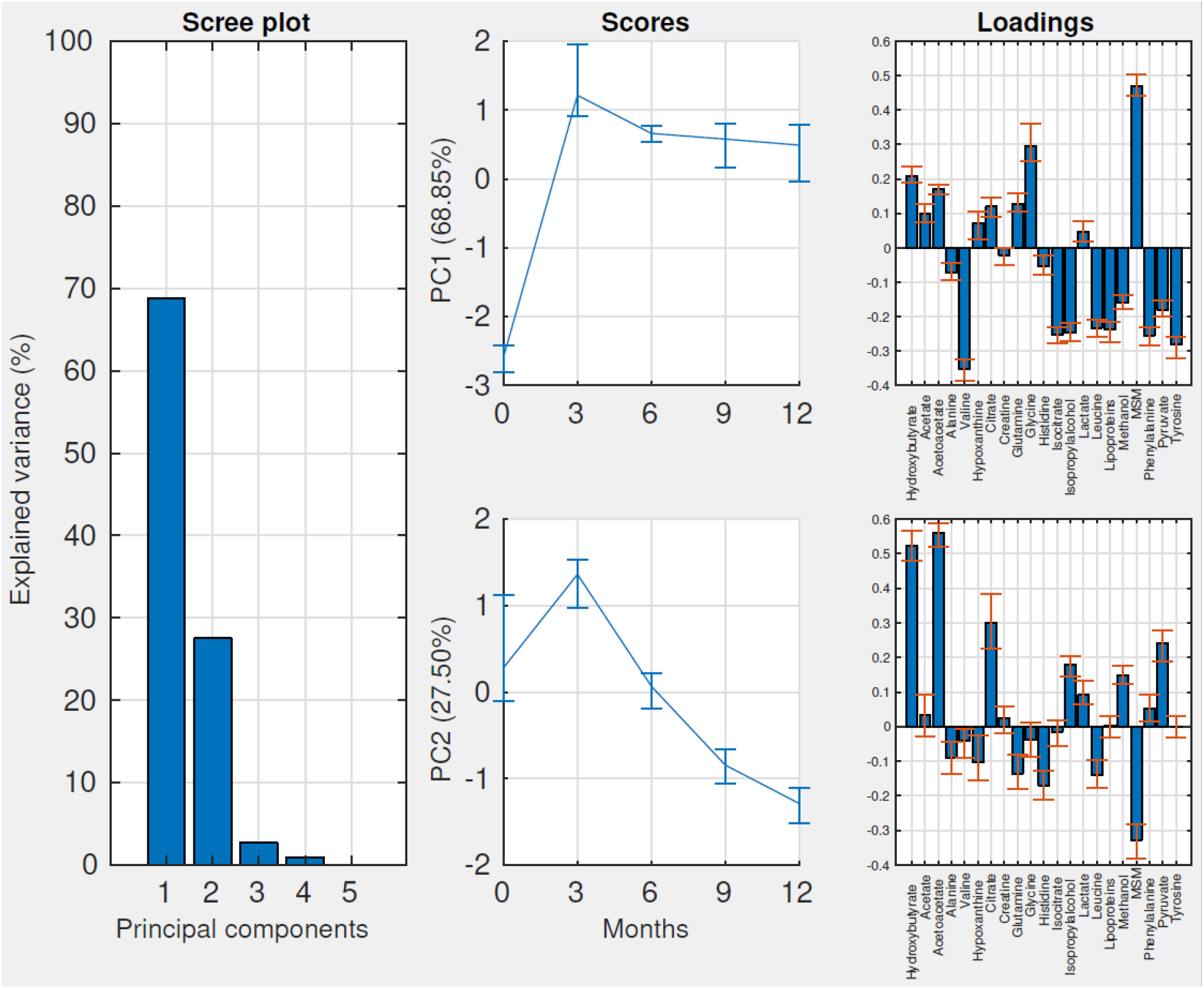
Scree-, score-, and loading plots for the effect matrix for time in the bariatric surgery data. Abbreviations: PC: principal component, MSM: methylsulfonylmethane.

#### 4.2.1 Time effect

The results from PCA on the effect matrix for time is shown in Figure 3. This trend represents the average change over time across all three groups. Two distinct patterns are observed in the score plots. Along the first component, which explains 69 % of the variance in the Time effect, there is a pronounced increase in score value between the first and second timepoint, and this difference persists over time. Metabolites with positive loadings on PC1 include methylsulfonylmethane, and the amino acid glycine, while the amino acids valine, isoleucine, tyrosine, and phenylalanine, the alcohols isopropylalcohol and methanol, and the lipoprotein signal have negative loadings.

In the second component, which explains 28 % of the variance in the time effect, a different pattern is observed. There is a temporary increase in scores after surgery, and then a progressive decrease over time. Metabolites with positive loadings on PC2 mainly include citrate and the ketone bodies acetoacetate and hydroxybutyrate, while methylsulfonylmethane has the most negative loading value.

#### 4.2.2 Treatment and time*treatment interaction effect

The results from PCA on the effect matrix for the treatment + time*treatment interaction effects is shown in Figure 4. The first principal component explains 64 % of the variation in the effects. In the score plot for PC1, the group receiving distal RYGB shows increasing score values over time, and diverges from the two other surgery groups. The loading plot for PC1 is characterized by a highly positive loading for methylsulfonylmethane. The fact that the PC1-scores of the three groups is the same at baseline suggests that the main source of variation is change in methylsulfonylmethane over time, and that this difference is not present at baseline. The baseline differences are visible at the first timepoint in PC2, which explains 14 % of the variation, but then start to significantly overlap at later timepoints.

**Figure 4:**
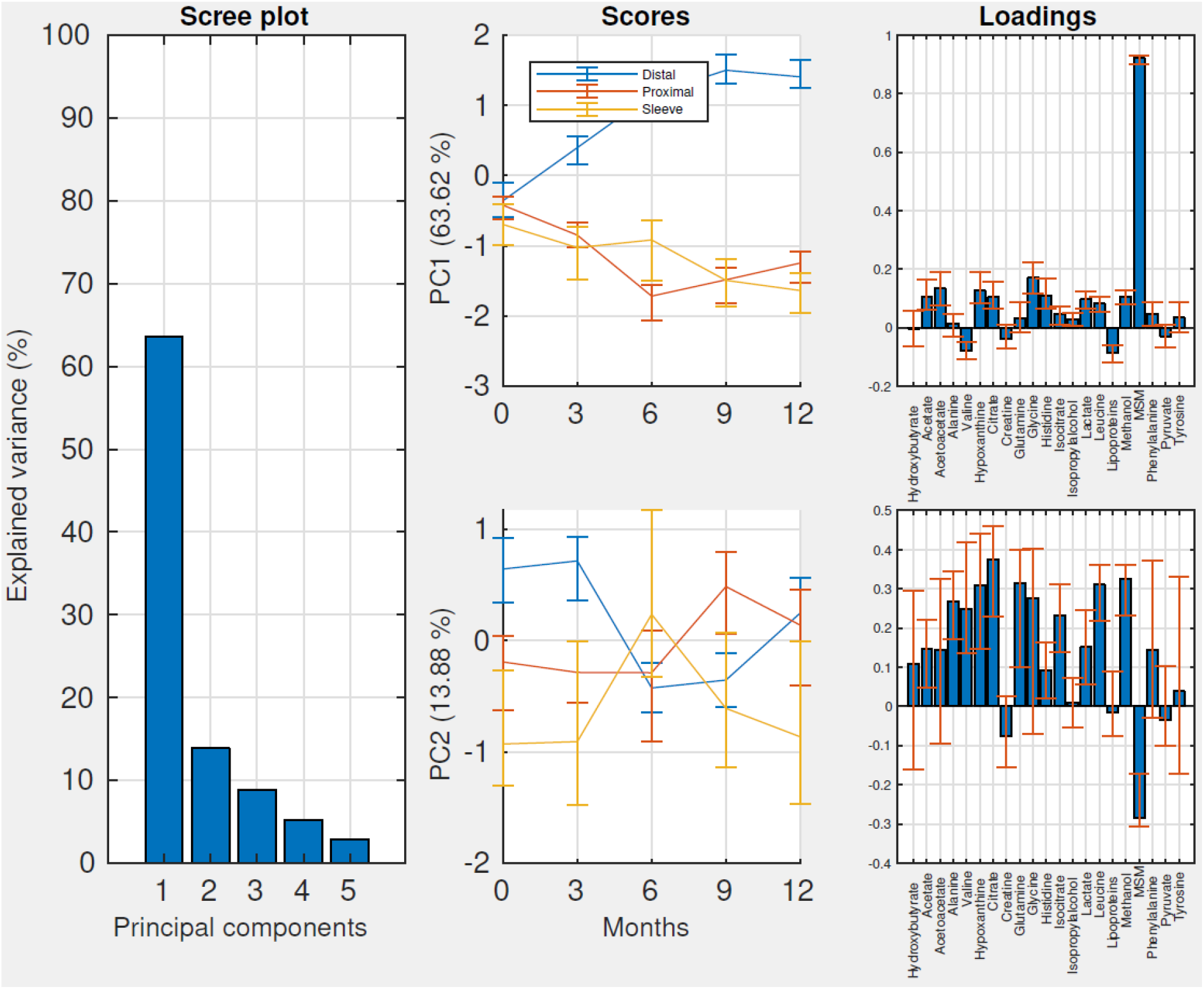
Scree-, score-, and loading plots for the effect matrix for the time*treatment interaction. Abbreviations: PC: principal component, MSM: methylsulfonylmethane.

## 5 Discussion

In this work we have described a novel methodology, RM-ASCA+, suitable for analysis of longitudinal multivariate data, and we have demonstrated this using two publicly available metabolomics datasets. We find that RM-ASCA+ yields interpretable and efficient representations of the findings in the original studies, while also revealing trends not previously apparent.

We have discussed three commonly used types of longitudinal linear models: 1) repeated measures models (cLDA/ucLDA), 2) longitudinal ANCOVA, and 3) analysis of changes. While the model types will in some settings yield equivalent results, and all will give unbiased estimates of the treatment effect for randomized studies (19), repeated measures models have some general advantages. In addition to allowing direct modeling of the time effect, which is useful when visualizing the effects over time in score plots, they can accommodate both randomized and non-randomized study designs. For these reasons, we find repeated measures models to be a suitable starting point for analyzing such studies using ASCA+.

For randomized studies, ucLDA and cLDA will usually yield similar score- and loading plots for the same dataset. However, in the case of chance imbalances in metabolites at baseline (which are always present), cLDA will adjust the effect estimates for regression to the mean (8). The intuitive explanation for this is that the group showing highest levels at baseline will on average be expected to decrease slightly at the next measurement, while the lowest group will be expected to increase. A consequence of this is that when estimating the treatment effect, an increase in the group showing the highest baseline levels should be given greater weight than an equivalent increase in the group with the lowest levels, because a further increase is more difficult to achieve if baseline levels are already high by chance. The same holds in the reverse situation. Models involving a baseline adjustment therefore give the most correct estimate of the treatment effect in this setting. The same does not hold for non-randomized studies, except for special situations where treatment assignment is determined by the baseline value (11). For most non-randomized studies, the treatment and control group will regress to different population means, and adjusting for baseline value of the response variable will introduce bias in the effect estimate. In such cases, limiting the analysis to comparison of within-group change may be more appropriate.

We have demonstrated RM-ASCA+ using data from two different studies. In the first study, by Euceda et al., the metabolic impact of neoadjuvant bevacizumab on tumor metabolism was assessed in a randomized controlled trial (16). The statistical analysis in the published paper involved a combination of PCA, PLS-DA, and univariate mixed models. A clear overall metabolic change over time for the entire cohort was described, characterized by increased levels of glucose and lipids, and decreased levels of phosphocholine, glycerophosphocholine, choline, and taurine, which was interpreted as signs of normalization of breast tissue metabolism (20). However, no significant discrimination was found at any timepoint between treatment and control by PLS-DA, and on univariate tests only glutathione was found to be significantly affected in the treatment group after adjustment for multiple tests. When applying RM-ASCA+, we find that the estimated scores of the treatment- and control group differ at all post-baseline timepoints, and the difference is robust to Jackknife-validation. The groups show directionally similar metabolic trajectories over time, but the slope is more steep for the treatment group. In PC2 we observe that both groups show a temporary increase in score value, which again is most pronounced in the group treated with bevacizumab, suggesting that the metabolites prominent in the loadings of PC2 (glutathione, choline, phosphocholine, glycine) are differently affected at this timepoint. This might reflect different chemotherapy regimens, as during the first half of the treatment the patients were treated with FEC (fluorouracil, epirubicine, and cyclophosphamide), while the remaining chemotherapy was taxane-based. Another possibility is that it is caused by discontinuation of active treatment, as patients were not given systemic cancer-directed therapies in the last four weeks before surgery.

In the second dataset included in this paper, Gralka et al. assessed metabolic changes in serum after bariatric surgery. They describe several changes, including increased levels of the amino acids glycine, glutamine, histidine, and arginine, along with increased levels of methylsulfonylmethane, trimethylamine-N-oxide, and formate, irrespective of procedure type. Conversely, concentrations of the branched chain amino acids (BCAA) isoleucine, leucine, valine, and the aromatic amino acids (AAA) phenylalanine and tyrosine were found to decrease, along with the lipoprotein signal and the gut microbiome derived metabolites methanol and isopropylalcohol. They also describe temporarily increased levels of the ketone bodies acetoacetate and hydroxybutyrate, and citrate after surgery, which was interpreted as reflective of ongoing fat catabolism. We find that by using RM-ASCA+ these results are visualized as two separate temporal trends. PC1 shows a large increase in score value from baseline to the first follow-up, and this score remains largely unaltered over time. This component describes the decreased levels of BCAAs, AAAs, lipoprotein signal, and methanol and isopropylalcohol, and increased levels of methylsulfonylmethane, glycine and ketone bodies. Both BCAAs and AAAs are implicated in the pathogenesis of obesity and metabolic syndrome, although their causal relationships remain unclear. Reduced serum levels of BCAAs can be observed after weight loss achieved by either surgery or diet, with the effect possibly being more pronounced for weight loss achieved through surgery (21-23). Like BCAAs, the AAAs phenylalanine and tyrosine have been positively associated with both prediabetes and T2D (24, 25), and changes during weight loss have generally been found to mirror changes in BCAAs (26). Glycine, which increased after bariatric surgery, has been inversely associated with metabolic syndrome, and positively associated with physical activity (27). In one study, reduced glycine levels in obesity was experimentally related to BCAA flux, where BCAA oversupply was hypothesized to interfere with oxidation of fatty acids, causing accumulation of lipid intermediates in the cell, which may deplete glycine levels through production of acylglycine (28). Increased glycine levels could therefore possibly be explained by the reduced supply of BCAAs.

The response pattern described by PC2 is characterized primarily by increased levels of ketone bodies and citrate. These metabolites increase after surgery, before decreasing over time and appearing to approach a steady state, which is not yet reached at the final follow-up. Both the temporal development and metabolite loadings of PC2 suggest that this component may reflect changes in fat catabolism, which presumably is highest in the first months after surgery, and then tapers off as the patients lose weight and reach a new energy equilibrium. Ketone bodies generally increase during periods of increased fat oxidation, due to increased availability of acetyl-CoA, which then react to produce ketone bodies (29). The explanation for increased citrate is less obvious, but levels of citrate, and other TCA-metabolites in general, have previously been reported to increase during prolonged fasting, possibly due to enhanced mitochondrial activity (30). As the prolonged caloric deficit after surgery may be metabolically similar to fasting, this could explain the increased citrate levels during the period of greatest weight loss.

Analysis of the time-treatment interaction effect showed that the metabolic effects of proximal RYGB and GS were similar, while the group receiving distal RYGB showed a different metabolic trajectory. This divergence was mainly driven by different effects on the metabolite methylsulfonylmethane. Methylsulfonylmethane increased in all surgery groups after surgery, but the group receiving distal RYGB showed more pronounced increases compared with proximal RYGB or GS. This difference was suggested by Gralka et al. to be due to the malabsorptive effects of distal RYGB, which could result in increased production of methylsulfonylmethane from the gut microbiome as a consequence of increased nutrient availability (18). Dietary supplementation of methylsulfonylmethane has been observed to improve the metabolic profile in obese mice (31). Moreover, observational studies suggest that malabsorptive procedures may produce superior improvements in blood pressure, lipid profile, and subsequent cardiovascular risk reduction, despite producing comparable weight loss (32, 33). These observations may imply that malabsorptive procedures may have additional benefits which are independent of their effects on weight loss. Other gut microbiome-derived metabolites observed to decline after surgery were methanol and isopropylalcohol. However, as these were equally decreased across all surgery types, they are likely altered by a different mechanism than methylsulfonylmethane.

While RM-ASCA+ can reveal interesting patterns in high-dimensional longitudinal data, some issues remain to be resolved. Although we have introduced ways of visually assessing the robustness of the results through jackknifing and augmenting the effect matrices with random effects and residuals, how to obtain p-values for the factor effects is not considered here. Another important issue is missing data. While LDA-models use all available data when estimating the effects, they require missing data to be missing at random (MAR) in order to provide unbiased estimates. This means that the probability of missingness must be conditionally independent of the value of the unobserved response, given the subject’s observed covariates (34). For example, if missing samples are metabolically different from non-missing samples from patients with the same covariates, missing data will bias the effect estimates regardless of which linear model is used. This is referred to as missing not at random (MNAR), or non-ignorable missing data. If there is substantial missing data, and it is suspected to be non-ignorable, it is recommended to perform multiple imputation to assess how different assumed distributions for the missing data affect the findings (35). A limitation of RM-ASCA+ compared to LiMM-PCA is that mixed models are computationally demanding, and applying mixed models to potentially tens of thousands of variables may not be feasible. In such situations, the pre-transformation of the response matrix by PCA done in LiMM-PCA can drastically reduce the number of response variables, making LiMM-PCA the more scalable alternative for high-dimensional data.

In conclusion, repeated measures linear mixed models can be used in conjunction with ASCA+ to visualize and compare multivariate changes between groups over time. This approach is not limited to metabolomics data, but may be suitable for any study using a longitudinal repeated measures design with a multivariate endpoint.

## Data Availability

The metabolomics-datasets used in this manuscript are publicly available, and can be found in the supplementary section at (https://doi.org/10.1007/s11306-017-1168-0), and on metabolomics data repository MetaboLights (MTBLS242). The data analysis code is available at (https://github.com/ntnu-mr-cancer/RM_ASCA)

https://www.ebi.ac.uk/metabolights/MTBLS242/descriptors

https://link.springer.com/article/10.1007/s11306-017-1168-0#Sec16

## 6 Acknowledgements

This work was supported by a grant from the Norwegian Research School in Bioinformatics, Biostatistics and Systems Biology (NORBIS), and a grant from the Norwegian Cancer Society.

The authors wish to acknowledge Leonardo Tenori (University of Florence) for providing information on the bariatric surgery study, and for publishing the metabolomics dataset. We would also like to acknowledge Tone F. Bathen (Norwegian University of Science and Technology) and Olav Engebråten (Oslo University Hospital) for publishing the metabolomics data from the NeoAva-study.

