## Supplementary Figures for "Repeated measures ASCA+ for analysis of longitudinal intervention studies with multivariate outcome data"

**Supplementary Figure 1 – Analysis of effect matrix for time (NeoAva)**

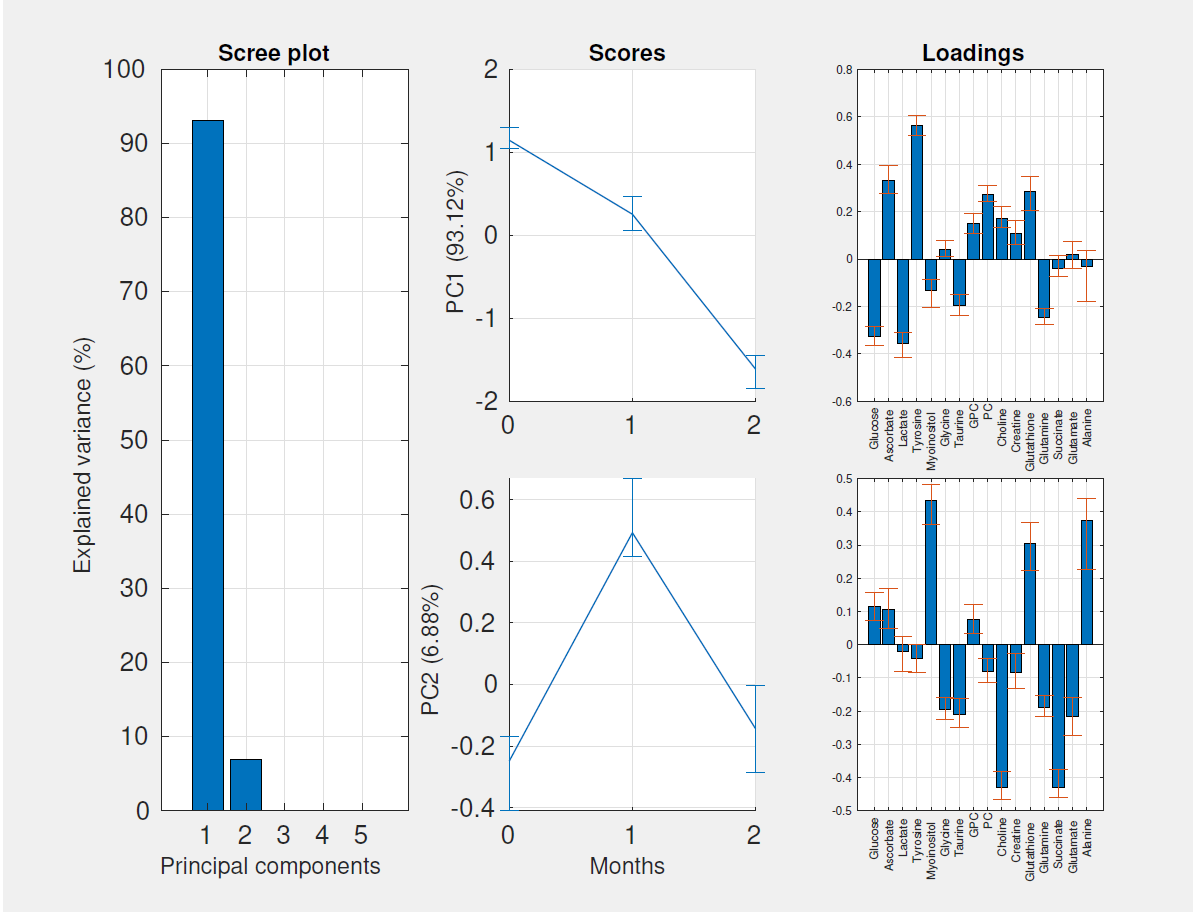

Supplementary Figure 1: Scree-, score-, and loading plots for the effect matrix for time for the NeoAva-data. Abbreviations: PC: Principal component, CTX: Chemotherapy, B: Bevacizumab, GPC: Glycerophosphocholine, PC: Phosphocholine.

**Supplementary Figure 2 – Analysis of effect matrix for treatment + time*treatment interaction (NeoAva)**

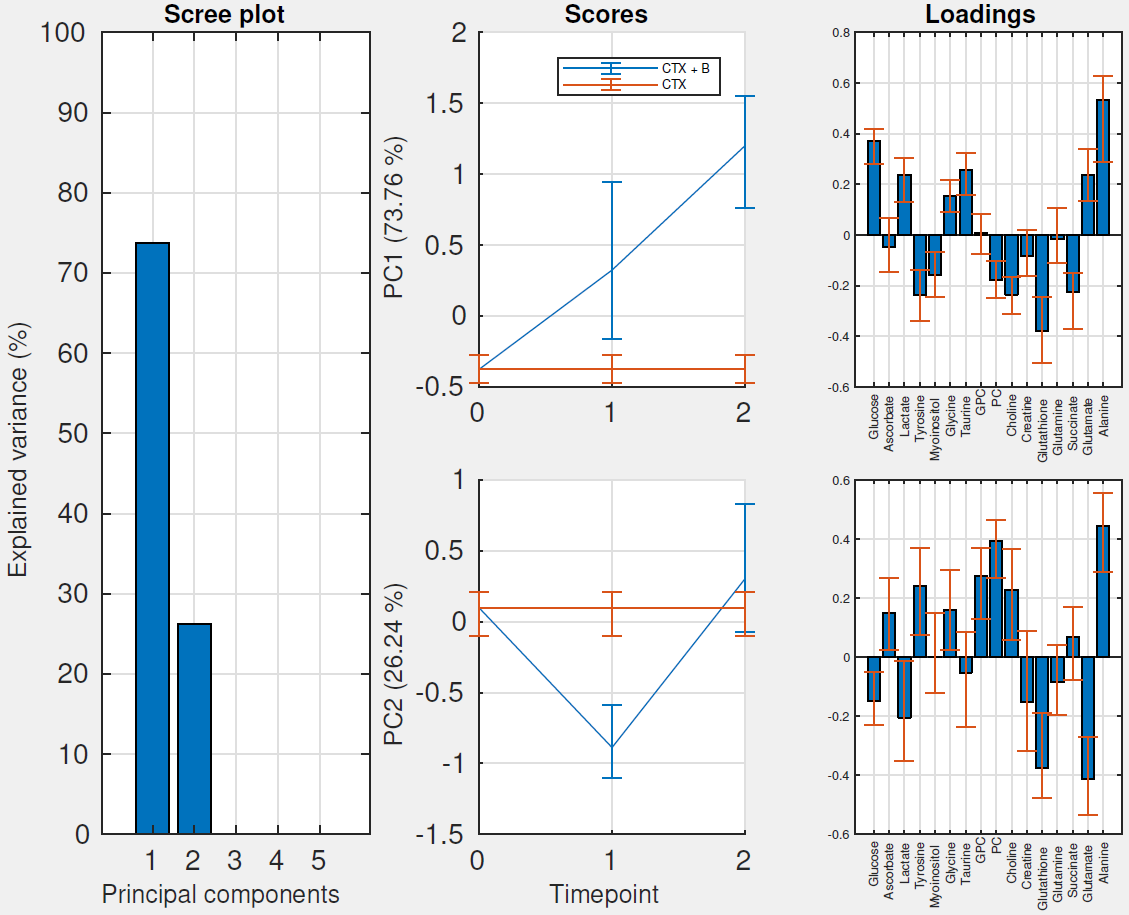

Supplementary Figure 2: Scree-, score-, and loading plots for the effect matrix for treatment + time*treatment interaction for the NeoAva-data. Abbreviations: PC: Principal component, CTX: Chemotherapy, B: Bevacizumab, GPC: Glycerophosphocholine, PC: Phosphocholine.

**Supplementary Figure 3 – Analysis of effect matrix for time + time*treatment interaction (Bariatric surgery data)**

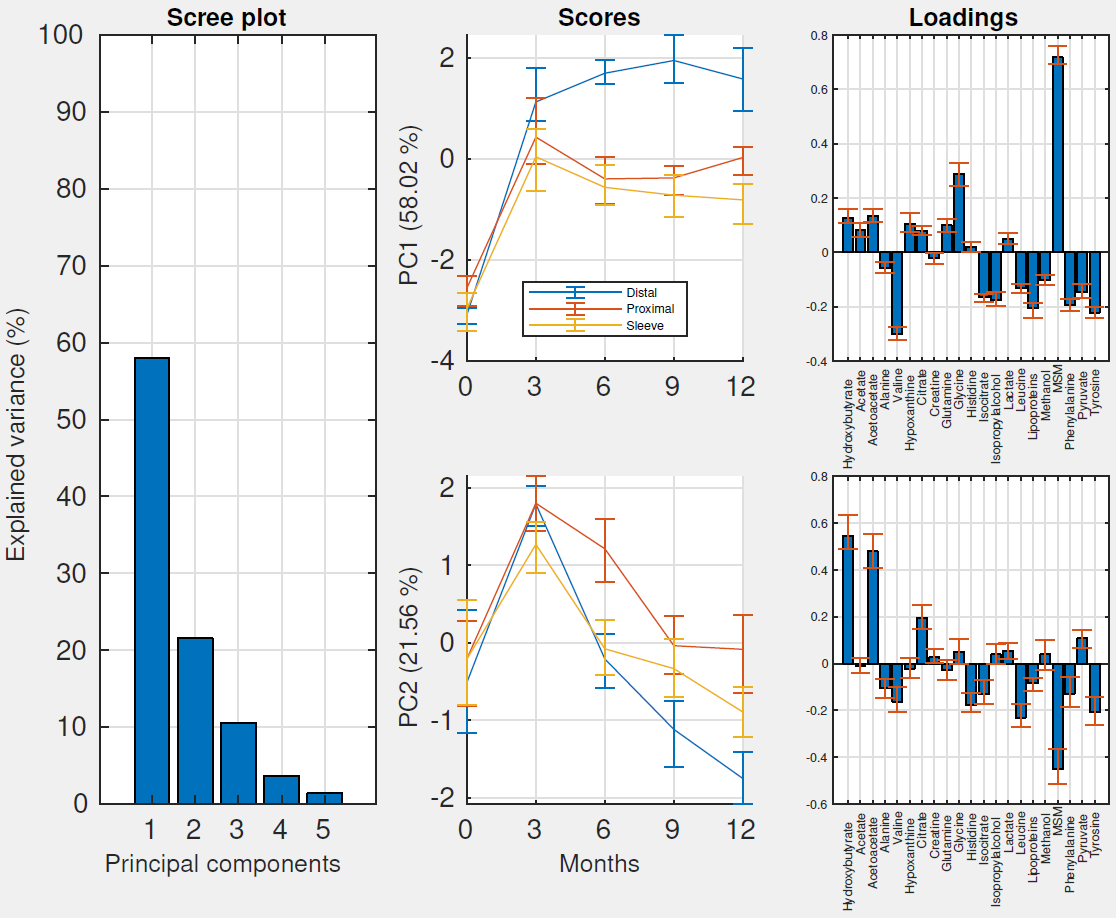

Supplementary Figure 3: Scree-, score-, and loading plots for the effect matrix for time + treatment + time*Treatment interaction for the bariatric surgery data. Abbreviations: PC: principal component, MSM: methylsulfonylmethane.
